# Averting an outbreak of Severe Acute Respiratory Syndrome Coronavirus 2 (SARS-CoV-2) in a university residence hall through wastewater surveillance

**DOI:** 10.1101/2021.06.23.21259176

**Authors:** Ryland Corchis-Scott, Qiudi Geng, Rajesh Seth, Rajan Ray, Mohsan Beg, Nihar Biswas, Lynn Charron, Kenneth D. Drouillard, Ramsey D’Souza, Daniel D. Heath, Chris Houser, Felicia Lawal, James McGinlay, Sherri Lynne Menard, Lisa A. Porter, Diane Rawlings, Yufeng Tong, Matthew L. Scholl, K.W. Michael Siu, Christopher G. Weisener, Steven. W. Wilhelm, R. Michael L. McKay

## Abstract

A wastewater surveillance program targeting a university residence hall was implemented during the spring semester 2021 as a proactive measure to avoid an outbreak of COVID-19 on campus. Over a period of 7 weeks from early February through late March 2021, wastewater originating from the residence hall was collected as grab samples 3 times per week. During this time, there was no detection of SARS-CoV-2 by RT-qPCR in the residence hall wastewater stream. Aiming to obtain a sample more representative of the residence hall community, a decision was made to use passive samplers beginning in late March onwards. Adopting a Moore Swab approach, SARS-CoV-2 was detected in wastewater samples on just two days after passive samplers were activated. These samples were also positive for the B.1.1.7 (Alpha) Variant of Concern (VOC) by RT-qPCR. The positive result triggered a public health case finding response including a mobile testing unit deployed to the residence hall the following day with testing of nearly 200 students and staff, which identified two laboratory-confirmed cases of B.1.1.7 variant COVID-19. These individuals were re-located to a separate quarantine facility averting an outbreak on campus. Aggregating wastewater and clinical data, the campus wastewater surveillance program has yielded the first estimates of fecal shedding rates of the B.1.1.7 VOC of SARS-CoV-2 in individuals from a non-clinical setting.

## Introduction

Novel coronavirus disease 2019 (COVID-19) is an acute respiratory disease that first came to the attention of the World Health Organization (WHO) in early 2020 (WHO 2020). The pathogen responsible for COVID-19 is SARS-CoV-2, a member of the coronavirus family (Coronaviridae Study Group 2020). By the end of 2020, >84 million cases and >1.8 million deaths had been reported world-wide (Worldometer 2020). These statistics, however, underestimate actual levels of infection as many patients are asymptomatic (Nishiura et al., 2020; Yanes-Lane et al., 2020; Oran and Topol, 2021) or present with only mild symptoms and do not seek medical attention. Indeed, undocumented infections may explain the initial rapid geographic spread of COVID-19 across the globe (Li et al., 2020). Therefore, it is of high priority to public health to optimize and expand appropriate screening and surveillance that can recognize the true prevalence of infection.

Wastewater monitoring offers a promising and cost-effective alternative to the large-scale testing of individuals as shown by a growing body of research from across the globe that has demonstrated the presence of SARS-CoV-2 RNA in sewage from wastewater treatment plants (WWTPs) (*e*.*g*., Ahmed et al., 2020a; Mallapaty, 2020, Medema et al., 2020), consistent with studies showing that the novel coronavirus is shed in feces (Wang et al., 2020, Wu et al., 2020). A 24-h composite sample of raw sewage represents the fecal discharge of the entire community served by the WWTP, effectively providing a community-wide swab. Taking into consideration rates of viral shedding, decay of the RNA signal within the sewershed, and sensitivity of the assay, modeling and numerical analysis can predict the ability to detect a single infection in a population from one hundred to 2 million people (Hart and Halden, 2020). Thus, data on SARS-CoV-2 viral load in wastewater can be used to inform municipalities and public health units on trends in community infections in the absence of widescale testing of individuals. This is consistent with wastewater-based epidemiology (WBE) programs implemented for pathogens such as the polio virus (Asghar et al., 2014, Mao et al., 2020). That SARS-CoV-2 can be shed even before manifestations of COVID-19 become apparent in an infected individual makes this approach even more powerful, especially for early warning (Jones et al., 2020). Indeed, WBE data from around the globe has identified SARS-CoV-2 RNA in wastewater prior to clinical metrics of infections in the communities served by those WWTPs (Medema et al., 2020, Peccia et al., 2020; D’Aoust et al., 2021a; Karthikeyan et al., 2021).

While informative of higher-level trends in community health (*e*.*g*., Wu et al., 2020), relying solely on WWTPs for sampling limits the epidemiological value of wastewater surveillance. Indeed, to yield most benefit, strategic sampling within a sewershed targeting neighbourhoods, schools or congregate living facilities provides finer spatial resolution that can result in actionable public health responses to limit or halt COVID-19 transmission (Hassard et al., 2021). Amongst early adopters of this approach have been colleges and universities throughout North America, many of whom are using WBE to monitor residence halls for early evidence of COVID-19 infection as an integral component of campus screening programs (Liu et al., 2020; Barich and Slonczewski, 2021; Betancourt et al., 2021; Bivins et al., 2021; Colosi et al., 2021; Gibas et al., 2021; Harris-Lovett et al., 2021; Karthikeyan et al., 2021; Reeves et al., 2021; Scott et al., 2021; Travis et al., 2021). In fact, by the completion of the 2020-21 academic year, >200 postsecondary institutions in North America, and >250 worldwide were involved in some form of wastewater surveillance on campus (Naughton et al., 2021). Yet, while there have been numerous examples where wastewater monitoring on a university campus has detected evidence for infection amongst community members, there are few examples where this monitoring triggered a public-health response that may have averted an actual outbreak (Betancourt et al., 2021; Gibas et al., 2021). Notable in this respect was a high-profile case at the University of Arizona in August 2020, where WBE triggered targeted clinical testing of students living in a residence hall that identified 3 individuals (2 of whom were asymptomatic) testing subsequently positive for COVID-19 (Betancourt et al., 2021; Schmitz et al., 2021). The infected students were relocated to a quarantine facility until they were deemed to be no longer infectious.

As part of the Province of Ontario’s Wastewater Surveillance Initiative (Government of Ontario, 2021), WBE was established at the University of Windsor, where wastewater originating from a residence hall was monitored thrice-weekly beginning in February 2021. Here we describe a case-study where wastewater surveillance triggered a public-health response that potentially averted an outbreak of the B.1.1.7 Variant of Concern (VOC) on a university campus.

## Materials and Methods

### Sample collection and location

The University of Windsor is a comprehensive public research university located in southwestern Ontario on the Canada-U.S.A. border enrolling >16,000 students. As with postsecondary institutions across Canada, the University transitioned to remote learning for the 2020-21 academic year (University Affairs, 2021). This reduced the footprint of students and employees on campus by ∼70% and meant that those students requiring on-campus accommodations could be housed in a single residence hall with a second on-campus residence used as necessary as a quarantine facility. Sampling of a residence hall on the campus of the University was initiated in early February 2021 that targeted a sewer line originating from the residence which empties into the municipal sewer system for the City of Windsor, Ontario. During the spring semester 2021, the residence hall housed 186 students living in 2-bedroom suites, with each suite sharing a common toilet facility. The residence hall contains two wings, each with separate sewer lines. The sewer line chosen for sampling serviced the north wing of the building which housed 86 students. From early February through late March, wastewater was collected as grab samples 3 times each week, usually between 10:00-11:00 h local time, using 500 mL polypropylene bottles. Beginning in late March, a passive sampler approach was adopted with the use of a modified Moore Swab (Sikorski and Levine, 2020; Liu et al., 2020; Bivens et al., 2021a). Briefly, this approach used a feminine hygiene product (tampon) connected by fishing line to a magnetic carabiner attached to the inside rim of the sewer cover. The modified Moore Swabs were deployed in duplicate into the wastewater stream where they resided for ∼20 h prior to retrieval. Deployment lasted from mid-afternoon through late morning the following day. Once retrieved, the swabs were collected into sealable plastic bags and transported in a cooler to the laboratory for processing. Time elapsed between sample collection and their receipt in the laboratory was no longer than 30 min.

### Sample processing

For grab samples of raw wastewater, a particle-associated fraction was concentrated by filtration through 0.22 µm Sterivex cartridge filters (MilliporeSigma, Burlington, MA), followed by flash-freezing the filter in liquid nitrogen as described previously (Chik et al., 2021). The filtrate was collected into a sterile 50-mL centrifuge tube followed by addition of 0.05% (v/v) Tween 20 and concentrated using a Concentrating Pipette (CP) Select (InnovaPrep, Drexel, MO) with Ultrafiltration PS Hollow Fiber Concentrating tips following a custom protocol (InnovaPrep, 2021). The CP Select concentrated the filtrate to ∼300 µL followed by flash-freezing in liquid nitrogen. Upon transition to use of passive samplers, individual Moore Swabs were placed into the barrel of a disposable 50-mL syringe and the filtrate plunged into a sterile 50-mL centrifuge tube. Filtrate was concentrated by ultrafiltration as described above using the CP Select. RNA was extracted from filters and concentrated filtrate following manufacturer’s instructions using the AllPrep PowerViral DNA/RNA kit (Qiagen, Germantown, MD). Samples were not treated with DNase upon extraction.

### RT-qPCR

Assays for SARS-CoV-2 targeted regions of the nucleocapsid (N) gene using United States (U.S.) Centers for Disease Control and Prevention (CDC) primers and probes for the N1 region (Lu et al., 2020). Assay of the B.1.1.7 VOC targeted a region of the N gene containing D3L, a signature mutation diagnostic of B.1.1.7 (Graber et al., 2021). The Pepper Mild Mottle Virus (PMMoV), which like SARS-CoV-2 is a positive-sense, single-stranded RNA virus, was selected as a fecal indicator and was quantified using primers and probes described previously (Haramoto et al., 2013).

Reactions contained 5 µl of RNA template mixed with 10 µl of 2 × RT-qPCR master mix (Takyon TM Dry One-Step RT Probe MasterMix No Rox, Eurogentec, Liège, Belgium) and primers and probes in a final reaction volume of 20 µL. Due to repeated incidence of inhibition with wastewater samples, template was diluted 1:5 in all reactions. Technical triplicates were run for detection of gene targets. Thermal cycling was performed using a MA6000 qPCR thermocycler (Sansure Biotech, Changsha, China). RT was performed at 48 °C for 10 min, followed by polymerase activation at 95 °C for 3 min, and 50 cycles of denaturation, annealing/extension at 95 °C for 10 sec, then 60 °C for 45 sec, respectively. The EDX SARS-CoV-2 synthetic RNA standard (Exact Diagnostics, Fort Worth, TX, USA) was used to create a 5-point standard curve to quantify the N1 gene target whereas synthetic RNA containing the D3L mutation (AR-S SARS-CoV-2 RNA Control 14; Twist Bioscience, South San Francisco, CA) served as a positive control for B.1.1.7. For PMMoV, a sample pooled from multiple WWTPs in

SW Ontario and quantified by RT-Droplet Digital PCR was used to generate standard curves (D’Aoust et al., 2021b). No template controls yielded no amplification, and we report a limit of detection of 5 gene copies of N1 per reaction (≥95% probability of detection).

### Estimating fecal shedding rates

The approach described as part of the WBE program at the University of Arizona (Schmitz et al., 2021) was adopted to estimate fecal shedding rates with a few modifications. This approach combines data from wastewater surveillance and clinical testing targeting SARS-CoV-2 along with estimates of flow rates during the period when the passive samplers were deployed. Fecal shedding rates in units of gene copies per gram-feces (gc/g-feces) were estimated using the equation:

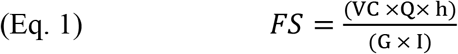

where, VC is the N1 gene concentration in units of gene copies per litre (gc/L), Q is the flow rate of wastewater leaving the residence hall in units of litres per minute and h is a conversion factor for time changing minutes to days. In the denominator, G represents the median per capita wet weight mass of feces from high income countries (126 g/person/day; Rose et al., 2015) and I is the total number of infected persons potentially contributing to the SARS-CoV-2 signal in wastewater.

Given challenges of determining absolute concentration of target genes using a passive sampler, VC was calculated from the ratio of SARS-CoV-2:PMMoV eluted from passive samplers and using the median PMMoV gene concentration determined from 17 grab samples collected over a 7-week period spanning February and March (Fig. 1). PMMoV is widely used as a fecal indicator to normalize SARS-CoV-2 in wastewater (Wu et al., 2020; D’Aoust et al., 2021b). Determining flow rates was likewise challenging given the small population serviced which generated intermittent flow through the sewers. Others have attempted direct measures of flow; however, problems exist using conventional flow meters especially when flow is very low and the sensors are not completely immersed in water (Schmitz et al., 2021). As an alternative, water consumption as measured by a public utilities meter in the building was used to estimate wastewater flow. Given that the water supplied to the student residence is used entirely within the building, the resulting wastewater flow discharged into the sewer system is expected to be very similar (Schilling and Tränckner, 2020). Water consumption was normalized to the number of students resident in the wing of the residence hall that was monitored and adjusted to better estimate percent water use during the normal time of deployment of passive samplers.

**Figure 1.**
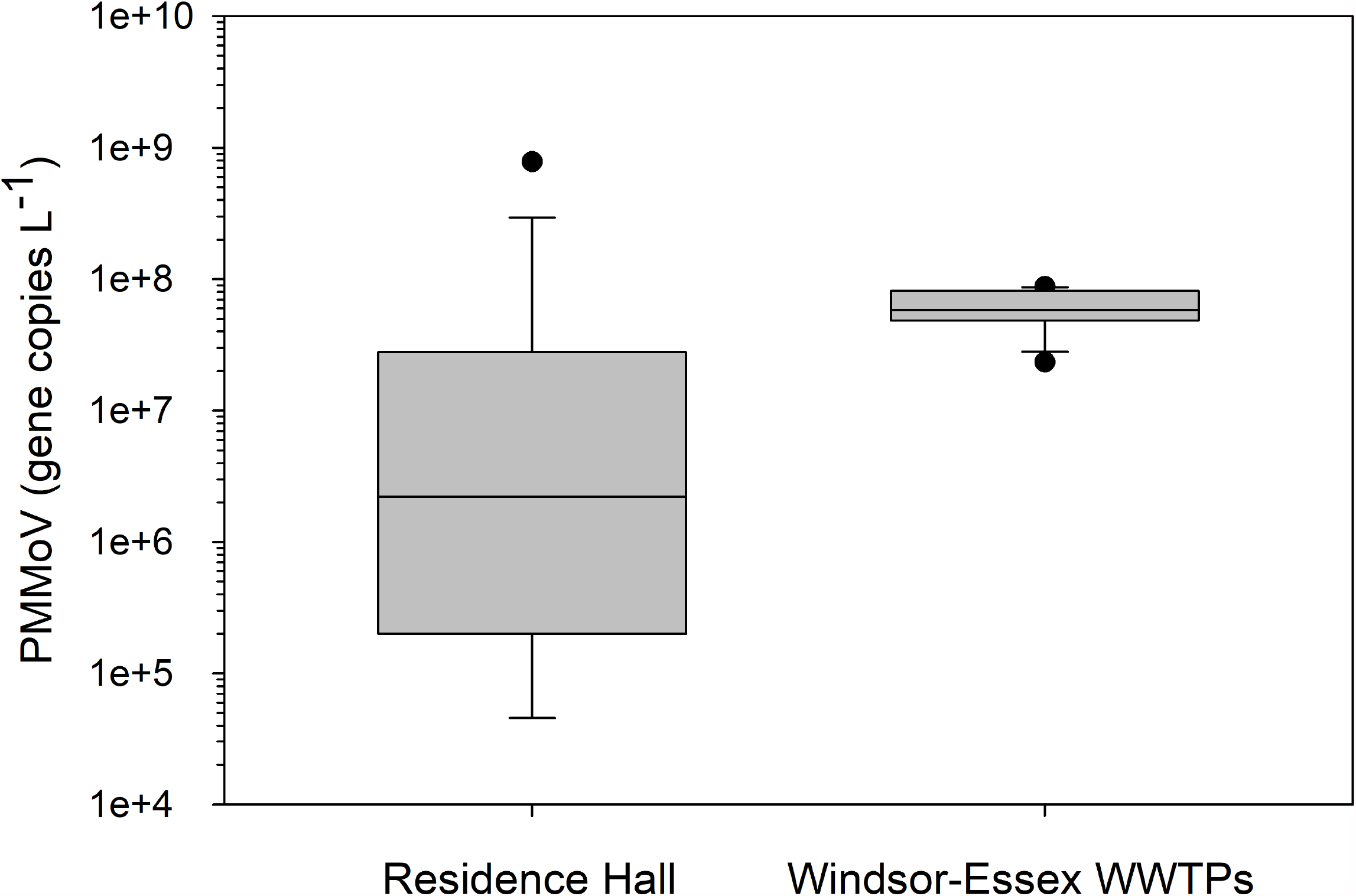
Concentration of PMMoV in wastewater from a university residence hall and as aggregate data across five WWTPs in Windsor-Essex County. Data are presented as box and whisker plots showing median gene concentrations. Vertical boxes around each median show the upper and lower quartiles whereas whiskers extend from the 10^th^ to 90^th^ percentile. Potential outliers are shown as discrete points.

## Results and Discussion

### Campus wastewater surveillance for SARS-CoV-2

A wastewater surveillance program on the campus of the University of Windsor was initiated in early February 2021, near the end of a Provincial lockdown as the Windsor-Essex County region was emerging from a resurgence of COVID-19 infections that spanned the months of December 2020 and January 2021 (Fig. 2). Within a week of initiating the program, restrictions were minimally relaxed as the region progressed to the Province of Ontario’s Red (Control) category, the second most restrictive category of the Province’s COVID-19 response framework (Government of Ontario, 2020).

**Figure 2.**
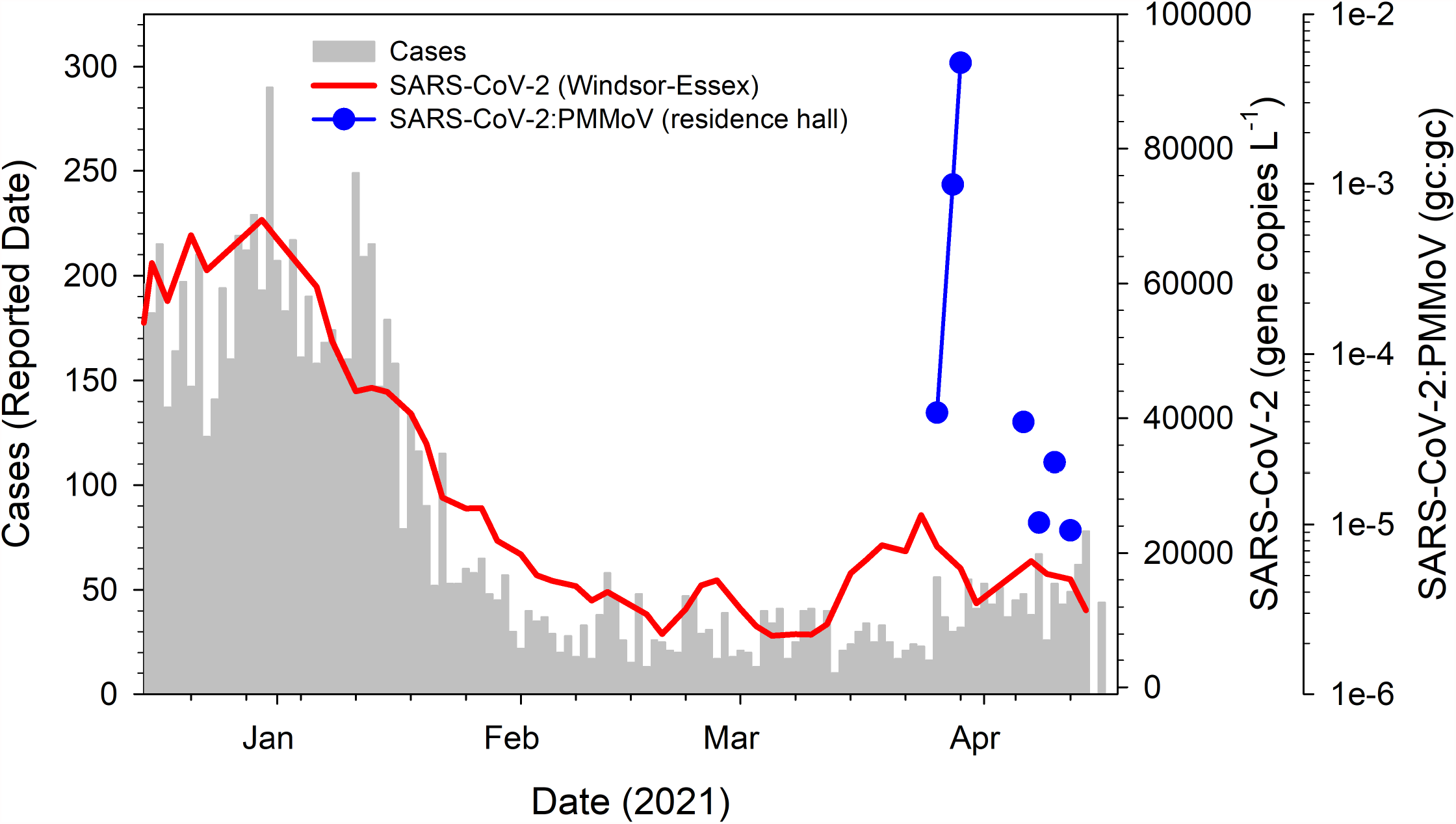
Concentration of SARS-CoV-2 N1 gene target in wastewater superimposed on COVID-19 cases in Windsor-Essex County plotted as a histogram. N1 gene concentration is a 7-day running average of aggregate data from five WWTPs in Windsor-Essex with data weighted by population served (red line). These data are publicly available on a dashboard updated weekly (WE-SPARK Health Institute, 2021). Following deployment of passive samplers, SARS-CoV-2 was detected in the residence hall sewer plotted as the ratio of gene copies (gc) of SARS-CoV-2:PMMoV (blue circles).

From early February through late March 2021, wastewater originating from a campus residence hall was collected as grab samples 3 times per week. Over this period, there was no detection of SARS-CoV-2 in the residence hall wastewater stream (data not shown). Over this same period, the concentration of SARS-CoV-2 in Windsor-Essex wastewater had stabilized at a low, but detectable level following the December-January resurgence of infections (Fig. 2).Comparability of data obtained from grab samples with 24-h composite samples obtained by autosampler has been investigated as part of several SARS-CoV-2 wastewater surveillance programs (Curtis et al., 2020; Bivens et al., 2021b; Gerrity et al., 2021). While general agreement between the approaches has been reported, groups report within-day variability in terms of detection of SARS-CoV-2 using the grab sample approach, a concern that is magnified when dealing with a congregate living facility housing a small population such as a university residence hall. This concern was reflected by the variability in the concentration of PMMoV yielded by grab samples which ranged across 4-orders of magnitude at the residence hall yielding a coefficient of variation (CV) of 2.83 (Fig. 1). In contrast, concentration of PMMoV from 24-h composite samples averaged across five WWTPs in Windsor-Essex over the same time showed only modest variation yielding a CV an order of magnitude lower at 0.38 (Fig. 1).

To obtain a sample more representative of the residence hall community and their defecation patterns (Heaton et al., 1992), a decision was made to implement the use of passive samplers from late March onwards. Unlike grab samples which represent a point-in-time ‘snapshot’, passive samplers offer the advantage of providing a time-integrated measure of the sampled matrix and potentially offers greater sensitivity of viral detection owing to the larger volume of sewage passing over the sampler compared to that which can be collected by a discrete sampling approach. Indeed, recent comparison of grab samples with Moore swabs targeting wastewater originating from a hospital admitting COVID-19 patients demonstrated that passive samplers can detect SARS-CoV-2 more consistently in wastewater (Liu et al., 2020). Likewise, Moore Swabs provided data comparable to an autosampler and both methods outperformed grab samples to detect SARS-CoV-2 from municipal sewer access points (Rafiee et al., 2021). Arguments against adopting the use of passive samplers include inability to quantify viral concentrations with confidence thus yielding a qualitative response. There are likewise uncertainties concerning the collection efficiency of the sampler, such as what fraction of virus passing across the sampler is retained, and is the virus retained by the sampler subject to degradation over the collection interval?

Moore Swabs yielded eluted concentrations of PMMoV that ranged over 2-orders of magnitude and a CV of 1.18 (data not shown). This was more variable than PMMoV yields obtained from composite samples from Windsor-Essex WWTPs over the same period but Moore Swabs offered improved consistency over grab samples based on this metric. SARS-CoV-2 was detected in wastewater eluted from Moore Swabs just two days after passive sampling was implemented (Fig. 2). The initial positive result triggered higher frequency sampling with passive samplers deployed daily the following week. Testing resulted in the detection of SARS-CoV-2 in the residence hall wastewater during the initial two days of daily sampling, following which the virus was no longer detected through the end of the week. During the initial 4-day period through which SARS-CoV-2 was detected, the signal continued to increase in intensity reflected by cycle threshold (Ct) values for the N1 gene target as high as ∼27. Normalizing to the fecal indicator PMMoV eluted from the same passive sampler, the ratio increased by 3-orders of magnitude over this 4-day period (Fig. 2). Following four days of negative tests, sampling resumed following a holiday weekend when SARS-CoV-2 was again detected in the residence hall wastewater although with signal intensity as normalized to PMMoV declining each day over a 7-day period.

At the time of the campus surveillance program, there was mounting concern globally over the emergence of SARS-CoV-2 VOCs. In North America, concern was focused largely on the B.1.1.7 VOC which was reported to be more transmissible than the wild-type Wuhan strain (Davies et al., 2021; Volz et al., 2021). Of particular concern to public health was a reported shift in the demographic of those infected trending to younger adults (Volz et al., 2021). Wastewater samples originating from the university residence hall were queried with an allele-specific primer extension RT-qPCR assay targeting a D3L mutation on the SARS-CoV-2 N gene that is diagnostic for B.1.1.7 (Graber et al., 2021). These samples tested positive for B.1.1.7 coincident with the initial detection of SARS-CoV-2 in residence hall wastewater in late March, and again in early April, following the reappearance of SARS-CoV-2 in the residence hall wastewater. While B.1.1.7 had become the dominant lineage of SARS-CoV-2 in parts of Ontario, especially in the Greater Toronto Area by mid-March (Brown et al., 2021), the Windsor-Essex region had reported just 26 cumulative cases assigned to B.1.1.7 by the time the VOC was detected in the residence hall wastewater (Windsor-Essex County Health Unit, 2021). Application of the allele-specific primer extension RT-qPCR assay to wastewater from Windsor-Essex showed no evidence of B.1.1.7 in the region through mid-March (R.M.L. McKay, unpublished), consistent with no more than 2 cases of B.1.1.7 reported in the region per day through our initial detection of SARS-CoV-2 in the residence hall waste stream (Windsor-Essex County Health Unit, 2021). Soon thereafter, this lineage had emerged with Windsor-Essex wastewater yielding a weighted mean of 14% B.1.1.7, which increased to ∼60% through weekly testing over the following month (WE-SPARK Health Institute, 2021). This B.1.1.7 signal rose in parallel with locations elsewhere in the province, albeit staggered in onset. The rapid increase in the dominance of B.1.1.7 in the wastewater was mirrored by clinical data with the region logging >1000 cumulative COVID-19 cases attributed to B.1.1.7 by early May (Ontario Agency for Health Protection and Promotion, 2021a).

### Insights into fecal shedding rates from wastewater surveillance

Since initial reports showing that SARS-CoV-2 can be detected in wastewater, a promising, albeit somewhat elusive extension of WBE, has been its use to estimate community infections within a sewer catchment (*e*.*g*., Ahmed et al., 2020; Curtis et al., 2020; Medema et al., 2020; Wu et al., 2020a; Chavarria-Miró et al., 2021). The uncertainties related to estimating absolute numbers of community infections are numerous and continue to be a challenge to realizing this application of WBE. Highlighting these uncertainties is our lack of understanding of fecal shedding rates as well as stability of the virus within the sewershed (Hart and Halden, 2020), where viral particles may be entrained from several hours to as long as ∼2 days depending on the sewer network (D’Aoust et al., 2021b).

Where prevalence of COVID-19 infections has been estimated from wastewater data, estimates of fecal shedding rates to derive loading of SARS-CoV-2 into wastewater have largely invoked data from a limited number of clinical studies examining excretion of virus in human feces. In these studies, the viral titer has been estimated to differ by several orders of magnitude (*e*.*g*., Wölfel et al., 2020; Zhang et al., 2020). Likewise, uncertainty surrounds the ubiquity and duration of fecal shedding. A systematic review and meta-analysis of 13 studies examining SARS-CoV-2 in stool showed mean shedding duration of 17.2 days with a maximum duration of 126 days (Cevik et al., 2020). Further, VOCs may exhibit different shedding patterns than the wild-type strain, with recent studies providing evidence for a higher viral load (Jones et al., 2021) and prolonged shedding time in the upper respiratory tract of patients infected with the B.1.1.7 lineage (Calistri et al., 2021; Kissler et al., 2021).

WBE programs associated with congregate living facilities offer a unique opportunity to calculate fecal shedding rates from a defined community. By combining wastewater surveillance with clinical data derived from testing individuals housed at these facilities, it is possible to extrapolate an approximate fecal shedding rate. Further, considering that sample collection points are typically adjacent to the facilities being tested, virus contributions to the wastewater stream are presumably recent thus negating some of the concerns over factors affecting the stability of SARS-CoV-2 in sewers. Aggregating wastewater and clinical data, the WBE program at the University of Arizona yielded amongst the first estimates of fecal shedding rates for SARS-CoV-2 from a non-clinical setting (Schmitz et al., 2021). In their study, implemented over a 3-month period in Fall 2020 and covering 13 dormitories, 81 wastewater samples tested positive for SARS-CoV-2 and triggered the clinical testing of students living in dorms resulting in diagnoses of 711 cases of COVID-19, of which 79.2% were classified as asymptomatic.

Because infected students were relocated to quarantine facilities that did not contribute to the study’s sewershed, infections associated with the dormitories were considered incident infections. Aggregating data from all dorms yielded a mean SARS-CoV-2 shedding rate of 6.84 ± 0.77 log10 gene copies/g-feces based on the N1 gene (Schmitz et al., 2021). The case study presented here based on the experience of implementing WBE at the University of Windsor likewise offered a unique opportunity to estimate fecal shedding rates attributed to a defined community, but with a signal easier to interpret compared with similar studies elsewhere. In this case, there was but a single occupied residence hall, having no detection of SARS-CoV-2 in wastewater over 7 weeks leading up to the initial detection, which triggered clinical testing of the building occupants. Two individuals tested positive for COVID-19 with both individuals relocated to a quarantine facility on campus less than two days after the wastewater data were reported to campus administration. Analysis of wastewater was positive for the B.1.1.7 VOC. Limiting our analysis to just the 4-day period encompassing the initial detection of SARS-CoV-2 in the residence hall wastewater, through to the clinical testing and quarantine of the two individuals who tested positive (Fig. 2), we report fecal shedding rates progressing in their intensity and ranging across 3-orders of magnitude from 3.93 log10 gc/g-feces to 5.99 log10 gc/g-feces based on the N1 gene target. These rates are lower than those reported from the Arizona study and must be interpreted with some caution, given the uncertainties surrounding fecal shedding, including reports that some infected individuals do not shed SARS-CoV-2 in their feces (Gupta et al., 2020; Jones et al., 2020). One must also consider the distinction that the cases presented here are specific to the B.1.1.7 VOC. Finally, we recognize that our estimates of fecal shedding rates are based on indirect assessment derived from ratios of SARS-CoV-2:PMMoV obtained from passive samplers, and using flow rates indirectly estimated from building water usage.

Despite uncertainty in estimating fecal shedding rates in this study, the linear progression in intensity of shedding over 4 days, as shown by the ratio SARS-CoV-2:PMMoV, was consistent with recent reports showing an estimated time from onset of shedding to peak viral load of 4.31 days (Jones et al., 2021) to around 6 days (Cavany et al., 2021). While we do not know if the subjects reached peak viral load by the time they were relocated to quarantine, the rapid increase in shedding intensity of more than 2000% as infection progressed between days 3 and 4 would suggest the peak was being approached.

Unfortunately, as sampling resumed following a holiday weekend and SARS-CoV-2 was once again detected in the residence hall wastewater, data interpretation was complicated due to the return of the students who had previously been quarantined. Thus, while a new infection was most likely responsible for the reemergent SARS-CoV-2 signal in wastewater detected in early April, that individual was removed to quarantine the following day and the 60% decline in signal that followed was attributed to reduced rates of shedding by the students deemed no longer to be infectious, but still classified as convalescent. A lack of detection of SARS-CoV-2 by mid-April corresponded with day 19 of the onset of infection associated with the initial subjects and thus close to the 17.2 day mean shedding duration reported previously (Cevik et al., 2020).

### Public health response and clinical confirmation of infection

Detection of SARS-CoV-2 in the residence hall wastewater sample triggered a rapid public health response by the University of Windsor COVID-19 Case Response Team and the Windsor-Essex County Health Unit (WECHU). Results from the initial detection of SARS-CoV-2 in residence hall wastewater were available by 12:00 h the day following Moore Swab retrieval and communicated to University administration by 17:00 h. University administration requested clarification regarding the data and conferred with Health and Safety at the University at which time the concern was elevated and WECHU was contacted by 20:00 h. Shortly thereafter, student residents and employees whose duties included access to the residence hall were sent electronic notification of the likelihood of a positive SARS-CoV-2 case within the facility and were encouraged to self-isolate. They were also apprised of a clinical testing unit to be mobilized to the residence hall the following morning. Over the following two days, over 195 nasopharyngeal swabs were collected by a mobile testing team with test results communicated within 24 h. From the initial cohort tested, a single positive SARS-CoV-2 infection was confirmed. This individual along with a close contact who was also a resident of the facility were moved to a separate quarantine facility on campus by midday the following day once initial test results were obtained. This close contact also tested positive as a part of testing of the second cohort one day later. RT-qPCR assay of the clinical samples for these individuals yielded Ct values ≤35 which triggered subsequent screening for the diagnostic N501Y, and E484K VOC mutations associated with the spike (S) gene using a multiplex RT-qPCR assay (Ontario Agency for Health Protection and Promotion, 2021b). Samples from both individuals were positive for the N501Y mutation and negative for E484K and presumed to be infected by the B.1.1.7 VOC based on Public Health Ontario criteria (Ontario Agency for Health Protection and Promotion, 2021b). This clinical diagnosis was consistent with wastewater testing which identified B.1.1.7 by targeting the diagnostic D3L mutation on the N-gene.

All other clinical tests performed following initial detection of SARS-CoV-2 in residence hall wastewater were negative. Quarantine of the two B.1.1.7-infected individuals had immediate implications for wastewater screening with a return to non-detects for SAR-CoV-2 during daily surveillance in the days following. With the resumption of testing following a holiday weekend yielding a positive result for SARS-CoV-2, the University COVID-19 Case Response Team was again notified, and we learned that a third student resident of the facility had been confirmed positive for COVID-19 earlier in the day after the student had sought testing the day prior. Upon learning of the test result, this third individual was relocated to the quarantine facility. Wastewater testing resumed the following day with another positive result suggesting that there remained one or more individuals at the residence hall actively shedding virus. Again, this result triggered a public health response and students were notified of a previously scheduled University-coordinated testing clinic on campus which attracted 65 student residents for testing, all of whom tested negative. We subsequently learned that the two residents who were initially quarantined had been approved to return to the residence hall after 10 days had elapsed, as they were deemed no longer infectious. This information combined with the knowledge that all student residents who chose to be tested following the re-emergence of SARS-CoV-2 in residence hall wastewater were negative suggests that convalescent shedding of SARS-CoV-2 was likely responsible for the virus persisting in the wastewater stream through mid-April.

## Conclusions

The University of Windsor has implemented a multi-pronged surveillance-based informative framework that combines wastewater testing with voluntary pooled saliva-based RT-qPCR screening to monitor for SARS-CoV-2 as part of a return to campus strategy (WE-SPARK Health Institute, 2021) that will continue into the Fall 2021 semester. Here we report on the wastewater testing component of this screening program and demonstrate that a WBE program targeting a congregate living facility on a university campus can lead to actionable responses by the university administration and public health, having the potential to avert an outbreak of COVID-19. As the COVID-19 pandemic unfolds, wastewater surveillance continues to be refined and informed by our growing understanding of SARS-CoV-2, emergence of variants, and persistence of the virus in wastewater. Actioning this emerging discipline of WBE into a public health response requires buy-in from administrators and public health authorities, confidence of which is often gained slowly. However, as with much of the decision-making associated with the pandemic, decisions have had to be made quickly without the benefit of having all evidence in place. In this case, growing confidence in this evolving discipline resulted in rapid deployment of a mobile testing team that identified infected individuals and resulted in their relocation to quarantine to avert an outbreak.

This work highlighted some of the challenges faced by WBE in congregate living facilities, including unpredictable wastewater flow and the confounding effect of convalescent shedding on interpreting the SARS-CoV-2 signal in wastewater. While many universities proactively relocate infected students into quarantine facilities, these are not meant to be long-term displacements. Students who are deemed no longer infectious, but still classified as convalescing, are normally approved to return to their assigned accommodations. Convalescent shedding of SARS-CoV-2 can persist for weeks to months, thus universities planning to use WBE need to consider this as part of their quarantine plans (Colosi et al., 2021).

As the current pandemic winds down with vaccination efforts ramping up globally, the longer-term application of WBE on campuses and elsewhere will need to be considered (Gibas et al, 2021): A recent commentary argued that wastewater surveillance can ‘have a second act’ to inform vaccine uptake especially if applied upstream within a sewershed to target neighbourhoods or congregate living facilities (Smith et al., 2021). These data would inform a public health strategy to encourage and facilitate vaccination of residents in these areas. Likewise, the value of WBE beyond the current pandemic is increasingly being recognized and there are calls to establish national wastewater surveillance systems having applications to detecting other well-known disease agents including foodborne pathogens shed in feces or to be applied to new pandemics caused by emerging pathogens (Keshaviah et al., 2021). Such calls to action are prompted by the rapid evolution of this public health tool as applied to COVID-19, and the realization that WBE represents a ‘community swab’ which is both cost-effective and scalable.

## Data Availability

Data available through online dashboard and Windsor-Essex County Health unit website.

https://www.wesparkhealth.com/covid-screening-platform#Dashboard

https://www.wechu.org/cv/local-updates

## Acknowledgements

We thank the Windsor-Essex County Health Unit’s mobile response team for its dedication and quick actions following alert by the University. We also thank J. Gauthier, L. Kiritsis, Z. Neale and V. Wert for providing support to the University of Windsor COVID-19 Case Response Team. We express appreciation to P. D’Aoust and R. Delatolla (University of Ottawa) for quantifying the PMMoV standard used in this project and to Y. Wu (S.M. Research, Inc.) who provided access to a thermal cycler. Funding in support of the Ontario Wastewater Surveillance Initiative was provided by the Ontario Ministry of Environment, Conservation and Parks. Additional support was provided by the Canada Foundation for Innovation - Exceptional Opportunities Fund (COVID-19 program) and by the Natural Sciences and Engineering Research Council of Canada (Alliance COVID-19 grant). Q. Geng received support through a MITACS Accelerate grant.

